# Southern European Prospective Investigation Into Childhood Cancer and Nutrition (EPIC*kids*): Study Design and Protocol

**DOI:** 10.1101/2025.01.29.25321322

**Authors:** Foteini Perganti, Inge Huybrechts, Adriana Cristina Balduzzi, Ronald Barr, Andrea Biondi, Aina Llenas Bladé, Evangelina Muñoz Bravo, Erika Damasco, Christina N. Katsagoni, Antonis Kattamis, Alvaro Lassaletta, Marta Llopis Lera, Anna Llort, Jessica Blanco Lopez, Antonio Perez Martinez, Maura Massimino, Andrés Morales La Madrid, Lucas Moreno, Ana Muñoz Alonso, Genevieve Nicolas, Giorgia Preziati, Sofia Rizzari, Serena Della Valle, Catalina Marquez Vega, Michelle Walters, Zisis Kozlakidis, Elena J. Ladas

## Abstract

The survival rates for children with cancer have increased appreciably over the last few decades; however, childhood cancer survivors continue to suffer from long-lasting sequelae. Studies have demonstrated that the presence of malnutrition, over- and under-nutrition, at diagnosis or the duration of malnutrition during treatment is associated with increased toxicity, infection, and inferior survival. Dietary habits, along with behavioral and socioeconomic status, are known factors that lead to obesity or undernutrition and can affect the prognosis and quality of life of children with cancer. Unfortunately, the underlying mechanisms responsible for these observations are largely unknown. To address this gap in science, we established the EPIC*kids* cohort study, an initiative of the International Initiative for Pediatrics and Nutrition at Columbia University Irving Medical Center and the International Agency for Research on Cancer of the World Health Organization. Over a 5-year period, children and adolescents with acute lymphoblastic leukemia and brain tumors receiving treatment in Spain, Italy, or Greece will be recruited. Clinical data and biospecimens (blood and stool) will be collected at designated timepoints in therapy. At the same time, several surveys will be administered to collect data on sociodemographics, physical activity, quality of life, food insecurity, and dietary habits. The primary aim of EPIC*kids* is to develop a large informative nutrition biobank and database to investigate the etiologic pathways that connect nutritional status and lifestyle factors with clinical outcomes in children and adolescents with cancer. Secondary aims are to create evidence-based guidelines for European children with cancer in this understudied region and to ultimately improve the quality of life of those children and adolescents.

The ClinicalTrials.gov ID for EPIC*kids* study is NCT05375617.

## Introduction

Every year, nearly 400,000 children and adolescents (0-19 years of age) are diagnosed with cancer worldwide. Leukemias and brain tumors are the most common types of cancer, with more than 80% of children with cancer cured in high-income countries (HICs). [1,2] Despite these accomplishments, the development of malnutrition (both over- and undernutrition) and nutrition-related conditions (e.g., metabolic syndrome, pancreatitis) predisposes children to adverse events during and after cancer care and negatively impact quality of life and survival.[3]

In children with acute lymphoblastic leukemia (ALL), several investigations have reported that poor nutritional status and dietary intake adversely impact prognosis and increase the incidence of several treatment-related toxicities.[4] Specifically, 5-year event-free survival was observed to be 64% and 65% for children with obesity or underweight at diagnosis, respectively, compared to 74% for those with a healthy weight (*P* = 0.002).[5] Obesity has consistently been associated with increased toxicities and inferior survival [6], which may be due to challenges with optimal drug dosing since increased adiposity may affect the pharmacokinetics and dynamics of lipophilic drugs.[7] Similarly, undernutrition is associated with numerous deleterious outcomes, such as reduced tolerance to treatment, increased toxicity, and infection.[3, 8–11] Up to 70% of children and adolescents with ALL develop overweight/obesity by the end of treatment in the United States.[12] Obesity has been a risk factor for metabolic syndrome [13], hyperglycemia [14], pancreatitis and hepatotoxicity in children with cancer.[6,15]

Children with brain tumors face similar challenges related to nutrition, although much less data is available. Undernutrition has been observed in 31% of children with medulloblastoma and other aggressive tumors at diagnosis [16], and it has been reported that up to 94% of children with medulloblastoma and 39% of children with other central nervous system tumors develop undernutrition during treatment.[17] Further, approximately 40% of children with brain tumors experience swallowing difficulties [18,19], which contribute to decreased oral intake and further decline in nutritional status. At the same time, the development of overweight/obesity is commonly observed in children and adolescents with hypothalamic damage due to radiation therapy, surgery, or by the tumor itself [20], resulting in endocrinopathies such as diabetes insipidus and central precocious puberty.[21] In one of the few reported European cohorts of survivors of a childhood brain tumor, a higher prevalence of overweight/obesity was observed as compared to the general population (29% compared to 13%, respectively) [21] as well as a greater percent of fat mass compared to non-cancer controls (mean difference 4.1%, 95% confidence interval 2.0-6.1).[22]

More recently, it has been reported that the duration of “aberrant nutritional status” (the time at which children are in an ‘at-risk’ nutrition state during treatment) appears to be more consistently associated with poor clinical outcomes.[23,24] However, the mechanisms by which nutrition impacts these outcomes are largely unknown.[25] Genetic predisposition [26,27], microbial environment [6] or parental feeding behaviors [28] may be responsible for excess weight gain. In children with ALL, remission induction therapy significantly reduces bacterial diversity in the gut and may contribute to the development of obesogenic microbial profiles.[29] The frequent use of antibiotic therapy during the induction phase of therapy causes additional insult to the intestinal microbiota, further augmenting this phenotype.[30] Unfortunately, much of the existing data are in children with ALL, with little information on the underlying mechanisms in children with brain tumors. Moreover, most findings are limited to children residing in North America.

Whether these findings are relevant to the European setting is unknown, given variations in treatment, lifestyle, environmental exposures, and the gut microbiome/metabolome. Studies have found that the Mediterranean diet and lifestyle are associated with reduced side effects of treatment, improved quality of life, and lower risk of adult-onset cancer.[31,32] The relevance in the pediatric setting is unknown. To address this gap in science, we established the Southern **E**uropean **P**rospective **I**nvestigation Into **C**hildhood Cancer and Nutrition (EPIC*kids*), which addresses this understudied area and establishes a novel infrastructure to comprehensively examine diet, lifestyle, and biological parameters among children and adolescents diagnosed with the two most common forms of childhood cancer.

## Material and methods

### Study design

The EPIC*kids* study (clinicaltrials.gov NCT05375617) is a prospective, observational, multinational cohort study recruiting children and adolescents with newly diagnosed ALL or favorable biology brain tumors and following participants from diagnosis until 1-year after the end of treatment (EOT). Recruitment began in June of 2023 and will continue through December 2028. Completion of the study is expected in January 2031, with analysis anticipated to be performed upon study completion. EPIC*kids* is being co-led by the International Initiative for Pediatrics and Nutrition (IIPAN), Columbia University Irving Medical Center (CUIMC) and the International Agency for Research on Cancer/World Health Organization (IARC/WHO).

Eligible participants are children and adolescents diagnosed with ALL meeting the following criteria: (1) aged between 3 and 21 years old at diagnosis with B-cell ALL, T-cell ALL, or mixed phenotype acute leukemia confirmed on immunophenotyping by flow cytometry and receiving standard leukemia treatment at one of the participating centers or may be children on a clinical trial or “as per” a clinical trial. For the brain tumor cohort, eligible children are: (1) aged between 3 years and 21 years old at the time of diagnosis with a favorable biology brain tumor confirmed by either pathology report, imaging and/or biochemical studies including low-grade gliomas, some medulloblastomas, ependymomas, pituitary tumors, germ cell tumors, and (2) receiving standard treatment at one of the participating centers with surgery, chemotherapy, radiation therapy or may be children on a clinical trial or “as per*”* a clinical trial. Children receiving autologous stem cell transplantation as part of their regimen may be included. Participating centers enrolling children and adolescents are listed in Table 1.

**Table 1.** PARTICIPATING CENTERS IN EPIC*kids* COHORT.

| **Table 1. PARTICIPATING CENTERS IN EPIC*kids* COHORT** |  |
| --- | --- |
| **Institution Name** | **Location** |
| Aghia Sofia Children’s Hospital | Athens, Greece |
| Fondazione IRCCS San Gerardo dei Tintori | Monza, Italy |
| Fondazione IRCCS Istituto Nazionale dei Tumori | Milan, Italy |
| Hospital Vall D’Hebrón | Barcelona, Spain |
| Pediatric Cancer Center Barcelona, Hospital Sant Joan de Déu | Barcelona, Spain |
| Hospital Infantil Universitario Niño Jesús | Madrid, Spain |
| Hospital Universitario La Paz | Madrid, Spain |
| Hospital Universitario Virgen del Rocío | Seville, Spain |

Each participating institution has a dedicated research assistant for the study, an institutional principal investigator (PI), and laboratory space for processing and storage of all stool and blood specimens. The biobank for all biological samples is centralized at IARC in Lyon, France, while the corresponding clinical data are managed by the IIPAN research team at CUIMC, New York, United States, using a REDCap database.

The primary aim of this study is to establish an informational resource on critical parameters of nutrition by which we can describe the trajectory of nutritional status among Southern European children and adolescents with ALL and favorable biology brain tumors, investigate lifestyle behaviors, sociodemographic factors, quality of life, and the intestinal microbiome/metabolome, and correlate these indicators with clinical outcomes.

### Clinical data collection

Initial and final risk groups and Grade 3 or 4 nutrition-related treatment-related toxicities (TRT) as defined by the National Cancer Institute Common Terminology Criteria for Adverse Events (CTCAE, version 5.0) will be abstracted from the medical records and input into the study’s REDCap database. The TRT that will be collected include all new cases of infection, mucositis, bone fractures, pancreatitis, hyperglycemia, liver toxicity, and peripheral neuropathy. Survival and clinical outcomes will be defined by uniform standards of practice in pediatric oncology: treatment-related mortality (TRM, death due to toxicity related to therapy), event-free survival (EFS, time from diagnosis to induction failure, relapse, secondary malignancy, or death), overall survival (OS, time from enrollment to death from any cause), and cumulative incidence of relapse (CIR, time to post-remission relapse). The number of days of hospitalization will also be collected. Administration of selected medications (e.g., steroids and antimicrobials) strongly associated with nutritional status or the microbiome will be collected prospectively. For participants receiving radiation therapy, information about the total dose and area/volume radiated will be collected. For the brain tumor cohort, information on pre-existing or newly developed endocrinopathies will be collected at each timepoint including growth hormone deficiency, pubertal delay, precocious puberty, hypogonadism, hypopituitarism, hypothyroidism, adrenocorticotropic hormone deficiency, and diabetes (type 2 diabetes mellitus or diabetes insipidus).

### Collection of biological specimens

An overview of the collection of biological specimens for the ALL and brain tumor cohorts is provided in Tables 2-3. For children with ALL, blood and stool will be collected at five timepoints (at diagnosis, at the end of induction, at the beginning of maintenance, at the end of treatment and 1 year after the end of therapy) representing times in which fluctuations in nutritional status have been reported. For the brain tumor cohort, the initial collection is after surgery, with further collections at 3-month intervals until one year after the end of treatment. Blood samples are obtained during a routine blood draw in serum separator tubes, centrifuged, and stored at -80°C until shipped for long-term storage at IARC. Stool samples are collected in the hospital or the home setting using a specified protocol. The collected stool is stored on fecal occult blood test cards and cryovials until shipped for long-term storage at IARC.

**Table 2.**
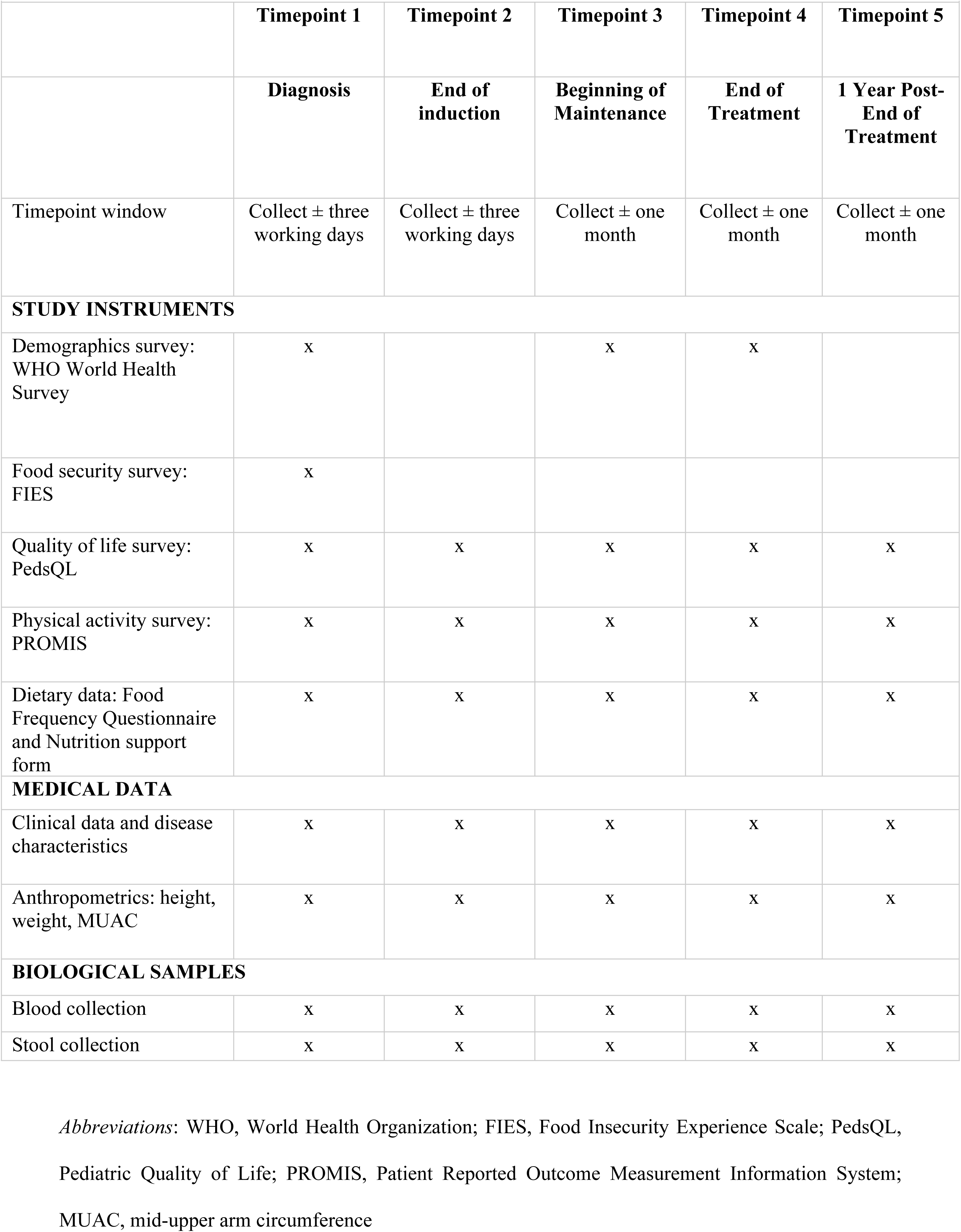
SUMMARY OF DATA COLLECTION: ACUTE LYMPHOBLASTIC LEUKEMIA COHORT.

|  |  |  |  |  |  |
| --- | --- | --- | --- | --- | --- |
|  | **Timepoint 1** | **Timepoint 2** | **Timepoint 3** | **Timepoint 4** | **Timepoint 5** |
|  | **Diagnosis** | **End of induction** | **Beginning of Maintenance** | **End of Treatment** | **1 Year Post-End of Treatment** |
| Timepoint window | Collect ± three working days | Collect ± three working days | Collect ± one month | Collect ± one month | Collect ± one month |
| **STUDY INSTRUMENTS** |  |  |  |  |  |
| Demographics survey: WHO World Health Survey | x |  | x | x |  |
| Food security survey: FIES | x |  |  |  |  |
| Quality of life survey: PedsQL | x | x | x | x | x |
| Physical activity survey: PROMIS | x | x | x | x | x |
| Dietary data: Food Frequency Questionnaire and Nutrition support form | x | x | x | x | x |
| **MEDICAL DATA** |  |  |  |  |  |
| Clinical data and disease characteristics | x | x | x | x | x |
| Anthropometrics: height, weight, MUAC | x | x | x | x | x |
| **BIOLOGICAL SAMPLES** |  |  |  |  |  |
| Blood collection | x | x | x | x | x |
| Stool collection | x | x | x | x | x |

**Table 3.**
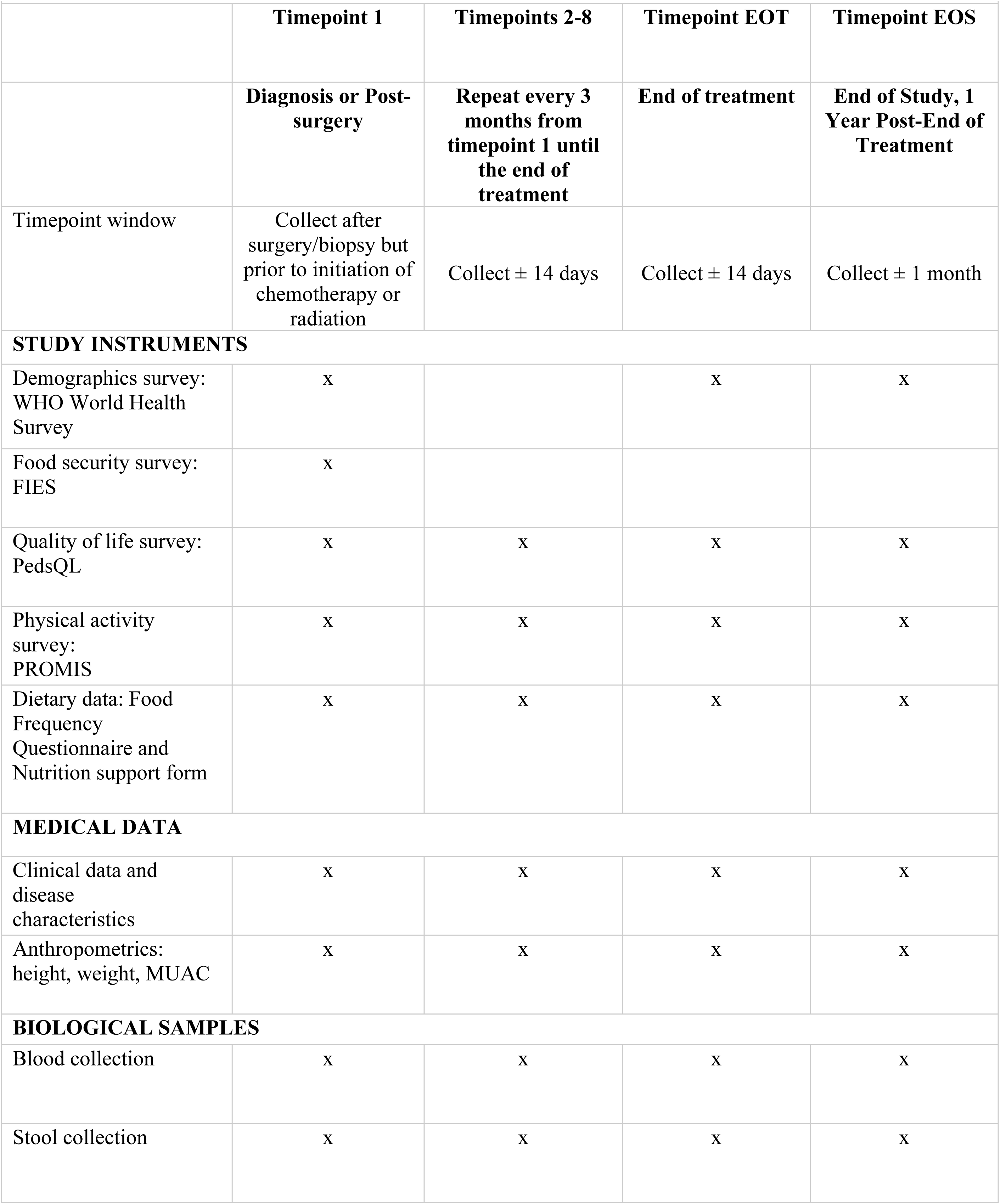
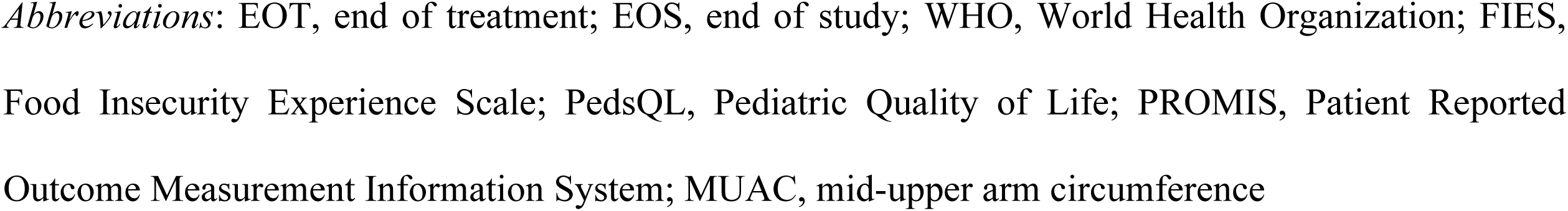
SUMMARY OF DATA COLLECTION: BRAIN TUMOR COHORT.

| **Table 3. SUMMARY OF DATA COLLECTION: BRAIN TUMOR COHORT** |  |  |  |  |
| --- | --- | --- | --- | --- |
|  | **Timepoint 1** | **Timepoints 2-8** | **Timepoint EOT** | **Timepoint EOS** |
|  | **Diagnosis or Post-surgery** | **Repeat every 3 months from timepoint 1 until the end of treatment** | **End of treatment** | **End of Study, 1 Year Post-End of Treatment** |
| Timepoint window | Collect after surgery/biopsy but prior to initiation of chemotherapy or radiation | Collect ± 14 days | Collect ± 14 days | Collect ± 1 month |
| **STUDY INSTRUMENTS** |  |  |  |  |
| Demographics survey: WHO World Health Survey | x |  | x | x |
| Food security survey: FIES | x |  |  |  |
| Quality of life survey: PedsQL | x | x | x | x |
| Physical activity survey: PROMIS | x | x | x | x |
| Dietary data: Food Frequency Questionnaire and Nutrition support form | x | x | x | x |
| **MEDICAL DATA** |  |  |  |  |
| Clinical data and disease characteristics | x | x | x | x |
| Anthropometrics: height, weight, MUAC | x | x | x | x |
| **BIOLOGICAL SAMPLES** |  |  |  |  |
| Blood collection | x | x | x | x |
| Stool collection | x | x | x | x |

*Abbreviations:* EOT, end of treatment; EOS, end of study; WHO, World Health Organization; FIES, Food Insecurity Experience Scale; PedsQL, Pediatric Quality of Life; PROMIS, Patient Reported Outcome Measurement Information System; MUAC, mid-upper arm circumference

### Survey Instruments

This study has five standardized surveys (Tables 2-3), either available in the native language of the country (English, Spanish, Italian, and Greek) or following appropriate forward/back translation and pilot tested among patients. The surveys are administered to the parent/participant by the local research personnel in their primary language. Questionnaire responses will be entered directly into the REDCap system and stored for analysis.[33,34] The REDCap database was selected as the electronic Case Report Form (CRF) platform because it is user-friendly, generates real-time reports, and data can easily be extracted for further analysis. Additionally, the REDCap mobile app’s utility [35] allows flexibility in administering the surveys to participants. A summary of the instruments is provided below.

#### Health-related Quality of Life

Health-related Quality of Life (HRQOL) is defined as the extent to which an individual’s usual or expected physical, emotional, and social well-being is affected by a medical condition or its treatment. The Pediatric Quality of Life (PedsQL) Generic Core Scales have been validated in pediatric oncology populations [36] and consist of 23 items, with a parent proxy report for ages 2-4 years, 5-7 years, 8-12 years, and 13-18 years and child self-report for ages 5-7 years, 8-12 years, and 13-21 years, yielding scores within the domains of Physical, Emotional, Social, and School Functioning.[37] The PedsQL survey responses are reverse-scored and transferred to a 0-100 scale. To reverse the score, the 0-4 responses will scale to 0-100: 0=100, 1=75, 2=50, 3=25, and 4=0. The scores are then averaged to a final scale score, computed as the sum of the items divided by the number of items answered, with higher scale scores indicating better HRQOL.

#### Food Insecurity Experience Scale (FIES)

The FIES [38] is a metric of the severity of food insecurity at the household or individual level and relies on direct yes/no responses to eight brief questions regarding access to adequate food. It is a statistical measurement scale similar to other widely accepted statistical scales designed to measure unobservable traits such as aptitude/ intelligence, personality, and a broad range of social, psychological, and health-related conditions. The order in which the eight FIES questions are asked follows the standard order survey module, in which questions refer to conditions that move roughly from less severe to more severe food insecurity. The FIES survey module is available in 200 languages, including those at all of the participating sites. The FIES survey module can be applied using different reference periods (30 days or 12 months). When the FIES is being applied to identify risk factors and consequences of food insecurity rather than for population monitoring, a 30-day reference period is recommended.

#### Physical Activity

The Patient Reported Outcome Measurement Information System (PROMIS) Pediatric Physical Activity survey is a validated, eight-item survey developed at the United States National Institutes of Health.[39,40] The survey assesses a child’s performance of activities that require physical actions which reflect the level of bodily movement, ranging from simple static behaviors with minimal muscle activity to more complex activities that require dynamic or sustained muscle activity and greater body movement. Physical activity behaviors are performed across non-hierarchical contexts related to the purpose (e.g., self-care, sports, school, recreation), physical environment (e.g., school, home, community), and social situations (e.g., alone, family, peers). Participants ≥8 years old will complete the self-report survey, and the parent will complete the parent-proxy survey. For participants ≤7 years old, parents will complete the parent-proxy survey on behalf of their child.

#### Demographics survey

Demographic data will be obtained utilizing an adapted version of the WHO’s World Health Survey (2002) emphasizing parental employment and education. [41]

#### Food Frequency Questionnaire (FFQ)

The detailed quantitative FFQ developed for this EPIC*kids* cohort study is based upon FFQs used in previous cohort studies investigating detailed dietary habits.[42] In addition to information regarding the frequency of consumption and the portion size usually consumed, this FFQ also includes extra specifications about the type of food usually consumed (e.g., fat or sugar content, cooking method, etc.).[42] The experience gained from the EPICSoft/GloboDiet methodology [43] implemented in many countries across the world was crucial to building the food lists and providing data and pictures (visual aids) used for estimating portion sizes.

The tool consists of approximately 100 food categories (e.g., root vegetables, poultry, cakes), grouped into 18 food groups (e.g., cereals, non-alcoholic beverages). This FFQ survey was standardized for all countries. The local, country-specific FFQs were generated by tailoring the FFQ to the local contexts and study population (e.g., including country-specific food lists). The FFQ is accompanied by a food image book featuring validated GloboDiet photos for foods and household measures (e.g., different sizes of a common glass) to aid participants in reporting actual intakes. The purpose of the FFQ is to obtain frequency and portion size information about food and beverage consumption over a designated period. Participants will be queried about their intake over the preceding month at each timepoint. To calculate the usual daily intake of foods, the frequency of consumption (in times per day) will be multiplied by the selected habitual portion size. Additional information regarding cooking method, source, fat/sugar content, etc., will be used to facilitate the matching with foods included in food composition tables to allow the computation of nutrient intakes.

### Study procedures

Approval was obtained from Human Research Protection Office and Institutional Review Board, CUIMC and the local IRBs at each participating site. Once written consent/assent (when applicable by age) is obtained by local PIs or delegated research personnel, each child is linked with a unique identification code in REDCap, which remains in a secure location at sites without access to external investigative staff. The delegated local dietician is responsible for completing the FFQ within the timeframe for each timepoint. During the designated timepoint window, blood and stool samples and anthropometrics (weight, height, mid-upper arm circumference) are obtained by the research staff, and. surveys are administered per the study protocol. All relevant clinical data are obtained from the medical chart and entered into the REDCap database.

### Data monitoring and quality control

#### Data Monitoring

Virtual data monitoring ensures completeness of data entry in REDCap and is performed following the designated timepoint windows. The local research personnel resolve any discrepancies unveiled by the monitoring report. Once data are verified, the CRFs are locked, and subsequent changes are not allowed. On site data monitoring occurs on an annual basis at which time data entry is verified with source documentation. Study IDs to be reviewed are generated by the study statistician. A written report is generated after each visit and any data concerns are discussed with the local PI and a remediation plan is established.

#### Quality Control and Assurance

Quarterly registration reports are generated to monitor accruals and completeness of registration data. Routine data quality reports are generated to assess missing data and inconsistencies. Accrual rates, accuracy of evaluations, and followup are monitored periodically throughout the study. Random-sample data quality and protocol compliance audits are conducted by the central study team at intervals outlined in the study protocol. The study committee (IARC, IIPAN, and participating centers) meets monthly to review the study progress report outlining accrual (including compliance with protocol enrollment criteria, reviewing the status of enrolled participants, dropouts, and losses to follow-up).

### Ethical and safety considerations

Consent and assent forms are translated into local languages, and the age limit for assent depends on local and national regulations. All study processes are performed according to the Good Clinical Practice, the Declaration of Helsinki, and the local principles of regulatory authorities. The personal data of children will be collected respecting the GDPR (General Data Protection Regulation). All study team members have been trained and are certified with the corresponding certificates prior to participation in the study.

Data confidentiality and privacy adhere to the policies outlined in the IARC Data Protection Policy Document for Scientific Processing. Medical information is confidential, and participants’ identities may not be used in written reports. Extensive measures will be taken to protect participant confidentiality. First, at the time a participant agrees to be included in the study, he/she is assigned a unique study ID. The study ID database containing the link between the unique identification number and participant name is maintained in a separate location from the REDCap database in a locked and secure area located at the participating institution. Access to the database in REDCap will be maintained by IIPAN at CUIMC.

Biospecimens bulk shipped to IARC will be stored with a code linked to each participant’s unique study ID. All information collected, including personally identifiable data, is protected using an advanced, multilayered security system based on standard authentication and authorization protocols. This includes server security, database security, and network security. All data are password protected and located on secure computers/servers, with limited access by investigators and staff involved in the study.

### Statistical analysis

The study protocol aims to describe the establishment of the infrastructure for this multinational collaboration. There are no statistical considerations for this study. Over five years, we propose to recruit 900 children and adolescents with newly diagnosed ALL and 1,400 children and adolescents with newly diagnosed brain tumors. This study will provide the opportunity for hypothesis-driven research proposals which will be submitted separately. Individual investigators are responsible for biostatistical planning in study design and outcome assessments according to the needs of specific projects.

Although this cohort study aims to collect clinical information and biospecimens from children undergoing cancer therapy to establish an information resource to address research questions yet to be defined, some basic analyses have been established a priori. We anticipate utilizing these data to estimate the power and sample size needed for future intervention studies, as well as answer some basic research questions such as: 1) How does dietary intake fluctuate during and after treatment for ALL or a brain tumor?; 2) What is the effect of treatment on nutritional status, defined by anthropometric measures (severely underweight, underweight, healthy weight, overweight, or obese) at baseline and at followup assessments? 3) What is the association between poor quality of life and nutritional status over the course of treatment? 4) What are the associations between food security, nutritional status (anthropometric), dietary intake, and treatment-related toxicities? 5) What are the overall trends of the aforementioned nutritional indicators and determinants (dietary intake, nutritional status, HRQOL, socioeconomic status) during the course of therapy? and 6) How are these nutritional indicators associated with survival in children with cancer? These analyses will allow the researchers and staff members to develop lifestyle interventions that may be tested through controlled, randomized trials to further improve outcomes among children with cancer.

### Current study status

Approvals from the IRBs have been obtained at all participating sites, and recruitment is ongoing in Greece, Spain, and Italy, with 72 children recruited thus far. Enrollment will remain open for a total period of five years.

## Discussion

The EPIC*kids* cohort study is a multinational study investigating children with ALL and brain tumors receiving treatment in Southern Europe; a group not well represented in nutrition and oncology studies. We anticipate the EPIC*kids* results will boost research studies to reveal the pathways connecting dietary habits, behavioral-socioeconomic status, biological pathways, and clinical outcomes. The results based on EPICkids data will be shared with the scientific community through national and international congresses, such as those organized by the International Society of Paediatric Oncology, the European Hematology Association, the Italian Association of Pediatric Hematology Oncology, the Spanish Association of Pediatric Hematology Oncology and the European Society for Paediatric Oncology in addition to investigator meetings and working groups relevant to the pediatric oncology field. Dissemination to the public will include media channels and articles in national and international press. Finally, we aim for the results based on EPIC*kids* data to be taken into consideration in public health actions, including meetings with national and international public health organizations. These efforts aim to foster the establishment of improved standards of care for children and adolescents with cancer in Europe. In parallel, educational seminars will be developed for oncologists, dieticians, nurses, patients, and parents.

## Data Availability

No pilot data are reported in the manuscript. Datasets were made only for FFQ verification. All relevant data from this study will be made available upon study completion.

## Acknowledgments

We thank the patients and their families, the physicians, dietitians, nurses, research coordinators, and all others who participated in preparing and collecting data for this cohort study.

## Disclaimer

Where authors are identified as personnel of the International Agency for Research on Cancer/WHO, the authors alone are responsible for the views expressed in this article and they do not necessarily represent the decisions, policy or views of the International Agency for Research on Cancer/WHO.

---

| SUMMARY OF DATA COLLECTION: BRAIN TUMOR COHORT |  |  |  |  |
| --- | --- | --- | --- | --- |
|  | Timepoint 1 | Timepoints 2-8 | Timepoint EOT | Timepoint EOS |
|  | Diagnosis or Post-surgery | Repeat every 3 months from timepoint 1 until the end of treatment | End of treatment | End of Study, 1 Year Post-End of Treatment |
| Timepoint window | Collect after surgery/biopsy but prior to initiation of chemotherapy or radiation | Collect ± 14 days | Collect ± 14 days | Collect ± 1 month |
| STUDY INSTRUMENTS |  |  |  |  |
| Demographics survey: WHO World Health Survey | x |  | x | x |
| Food security survey: FIES | x |  |  |  |
| Quality of life survey: PedsQL | x | x | x | x |
| Physical activity survey: PROMIS | x | x | x | x |
| Dietary data: Food Frequency Questionnaire and Nutrition support form | x | x | x | x |
| MEDICAL DATA |  |  |  |  |
| Clinical data and disease characteristics | x | x | x | x |
| Anthropometrics: height, weight, MUAC | x | x | x | x |
| BIOLOGICAL SAMPLES |  |  |  |  |
| Blood collection | x | x | x | x |
| Stool collection | x | x | x | x |

SPIRIT schedule for enrolment, interventions, and assessments -
| SUMMARY OF DATA COLLECTION: ACUTE LYMPHOBLASTIC LEUKEMIA COHORT |  |  |  |  |  |
| --- | --- | --- | --- | --- | --- |
|  | Timepoint 1 | Timepoint 2 | Timepoint 3 | Timepoint 4 | Timepoint 5 |
|  | Diagnosis | End of induction | Beginning of Maintenance | End of Treatment | 1 Year Post-End of Treatment |
| Timepoint window | Collect ± three working days | Collect ± three working days | Collect ± one month | Collect ± one month | Collect ± one month |
| STUDY INSTRUMENTS |  |  |  |  |  |
| Demographics survey: WHO World Health Survey | x |  | x | x |  |
| Food security survey: FIES | x |  |  |  |  |
| Quality of life survey: PedsQL | x | x | x | x | x |
| Physical activity survey: PROMIS | x | x | x | x | x |
| Dietary data: Food Frequency Questionnaire and Nutrition support form | x | x | x | x | x |
| MEDICAL DATA |  |  |  |  |  |
| Clinical data and disease characteristics | x | x | x | x | x |
| Anthropometrics: height, weight, MUAC | x | x | x | x | x |
| BIOLOGICAL SAMPLES |  |  |  |  |  |
| Blood collection | x | x | x | x | x |
| Stool collection | x | x | x | x | x |

